# Acoustic enrichment of heterogenous circulating tumor cells and clusters from patients with metastatic prostate cancer

**DOI:** 10.1101/2023.12.04.23299128

**Authors:** Cecilia Magnusson, Per Augustsson, Eva Undvall Anand, Andreas Lenshof, Andreas Josefsson, Karin Welén, Anders Bjartell, Yvonne Ceder, Hans Lilja, Thomas Laurell

## Abstract

**Background:** There are important unmet clinical needs to develop cell enrichment technologies to enable unbiased label-free isolation of both single cell and clusters of circulating tumor cells (CTCs) manifesting heterogeneous lineage specificity. Here, we report a pilot study based on microfluidic acoustophoresis enrichment of CTCs using the CellSearch CTC assay as a reference modality.

**Methods:** Acoustophoresis uses an ultrasonic standing wave field to separate cells based on biomechanical properties (size, density, and compressibility) resulting in inherently label-free and epitope-independent cell enrichment. Following red blood cell lysis and paraformaldehyde fixation, 6 mL of whole blood from 12 patients with metastatic prostate cancer and 20 healthy controls were processed with acoustophoresis and subsequent image cytometry.

**Results:** Acoustophoresis enabled enrichment and characterization of phenotypic CTCs (EpCAM^+^, Cytokeratin^+^, DAPI^+^, CD45^-^/CD66b^-^) in all patients with metastatic prostate cancer and detected CTC-clusters composed of only CTCs or heterogenous aggregates of CTCs clustered with various types of white blood cells in 9 out of 12 patients. By contrast, CellSearch did not detect any CTC-clusters, but detected comparable numbers of phenotypic CTCs as acoustophoresis, with trends of finding higher number of CTCs using acoustophoresis.

**Conclusion:** Our preliminary data indicate that acoustophoresis provides excellent possibilities to detect and characterize CTC-clusters as a putative marker of metastatic disease and outcomes. Moreover, acoustophoresis enables sensitive label-free enrichment of cells with epithelial phenotype in blood and offers opportunities to detect and characterize CTCs undergoing epithelial-to-mesenchymal transitioning and lineage plasticity.

---

There is an urgent need to develop pre- and post-treatment biomarkers for prostate cancer (PCa) to aid in clinical decision-making throughout the various disease stages. The idea to use liquid biopsy to monitor the disease progression for epithelial cancers motivates a strong interest in identifying methods to isolate and enumerate rare circulating tumor cells (CTCs). Several studies showed an association between higher counts of CTCs in blood from metastatic cancer patients to a poor prognosis and low overall survival (1, 2). Still today, the CellSearch CTC enumeration system is the only US Food and Drug administration (FDA) cleared CTC-technology to be used as a prognostic biomarker predictive of overall survival in epithelial cancers (1-4), and is therefore, commonly used as reference modality when validating novel CTC-technologies.

Like several other CTC targeting assays, the CellSearch system captures and identifies CTCs by the epithelial cell adhesion molecule (EpCAM), which is exclusively expressed in epithelia and in epithelial derived carcinomas. The major drawback with a positive selection approach is the inability to detect various subtypes of CTCs with reduced or absent EpCAM expression (5-8), e.g., due to an epithelial to mesenchymal transition (EMT). Such a transition is considered a pre-requisite for tumor cell infiltration and metastasis formation at secondary sites (9). Studies have found that patients with a poor response to chemotherapy had significantly more CTCs of mesenchymal-like phenotype compared to patients who responded to the treatment (10-12). Numerous studies have shown that CTCs also exist in cell clusters, although extremely rare and only representing a few percent of the total number of CTCs (13). Such clusters have previously been demonstrated to provide up to fifty-fold higher metastatic capacity than single CTCs (13). Notably, CTCs can also form clusters with white blood cells (WBCs) (14), which likely reduces the chance of capture in methods purely based on negative selection targeting CD45.

The emergence of high precision microfluidics for cell separation and sorting have resulted in new CTC-technologies based on different principles where a major category of methods rely on antibody capture to enrich CTCs, such as antibody-coated microstructures (15, 16), and magnetophoresis (17, 18). Other methods exploit differences in the biophysical properties of cells, such as cell-size-dependent deterministic lateral displacement (19), size-and-density-dependent inertial separation (20, 21), or electrical-conductivity-dependent dielectrophoresis (22, 23). Although more complex, hyphenation of several microfluidic techniques has proven to be beneficial in cell and CTC separation (15, 19, 24, 25).

The various methods for CTC enrichment usually result in a selection bias of the targeted cells, either by the epithelial marker expression level or cell size as these are the most common discriminators. Acoustophoresis has emerged as an alternative tumor cell separation technology. This method separates cells based on their acoustic mobility, for which the cell density, compressibility, and size are the determining factors (26). Acoustophoresis uses an ultrasonic standing wave field to manipulate cells in microfluidic channels and is inherently label- and contact-free and proven to be gentle to cells (27), which is important in detection of CTC-clusters (multicellular CTC-aggregates). Separation of human blood cells based on microchip acoustophoresis, i.e. free-flow acoustophoresis (FFA), which was first reported by Petersson et, al. (28). Augustsson et al. later refined the resolution of FFA, pioneering tumor cell isolation from WBCs (26). Magnusson et al. further demonstrated that the throughput of tumor cell acoustophoresis could be scaled to match clinical needs, separating a 5 mL spiked blood sample in 2 hours (29). Using an alternative acoustophoresis technique, i.e., tilted angle surface acoustic wave, Li et al. demonstrated that CTCs could be detected in two out of three breast cancer patients at modest flowrate and without reference to a healthy base line control (30). Acoustophoresis offers EpCAM-independent enrichment of CTCs, which enables detection of additional CTC subtypes with an EMT profile. We here address the next step in the analytical validation process of acoustophoresis and for the first time present acoustophoretic CTC and CTC-cluster isolation from clinical samples including baseline measurements from healthy controls and subsequently comparison versus CellSearch.

## METHODS

### Study assessments

The primary objective of the study was to assess performance characteristics of label-free microfluidic acoustophoresis CTC enrichment identified by phenotypic expression pattern (EpCAM^+^, CK^+^, CD45^-^, DAPI^+^) followed by morphological characteristics derived by image flow cytometry (IFC). The number of CTCs was compared to CellSearch. We also evaluated the ability of acoustophoresis to enrich subtypes of CTCs and CTC-clusters.

### Blood sample collection and study participants

Ethylenediaminetetraacetic acid (EDTA) anti-coagulated whole blood (6 mL) was collected in Vacutainer tubes (BD Bioscience, Temse, Belgium) at Skåne University Hospital (Malmö, Sweden) and Sahlgrenska University Hospital (Gothenburg, Sweden) from 12 men (aged 58-91 years) with metastatic prostate cancer (mPCa), ***see Supplemental Table 1***. The project was carried out in accordance with Helsinki declaration and approved by local ethical comities (Approval No. 367-03; 936-12), all participants gave informed consent. A concurrent collection of 7.5 mL of blood in CellSave Vacutainer tubes (Menarini Silicon Biosystems, Milan, Italy) obtained from 10/12 mPCa cases was used to compare the performance characteristics of acoustophoresis with the CellSearch assay. EDTA-anticoagulated blood (6 mL) was collected from anonymized healthy volunteers providing signed informed consent at the Biomedical Center, Lund University (Lund, Sweden) according to a protocol approved by the Swedish ethical review authority (ref. no. 2020-05818). Blood samples designated for acoustic separation were subjected to red blood cell (RBC) lysis and paraformaldehyde (PFA) fixation within 4 hrs. of venipuncture and stored in buffer-A [1x phosphate buffered saline (PBS), 1% fetal bovine serum (FBS) and 2 mM EDTA] at 4°C until processed.

#### Acoustophoretic setup

The CTC separation platform has been previously described (29). Briefly, an acoustofluidic microchip was manufactured in silicon and glass using standard microfabrication processing (31). The acoustofluidic chip, ***Fig. 1A***, has an initial pre-focusing channel (length × width × depth: 20 mm × 300 μm × 150 μm), in which the cells are exposed to a ∼5 MHz resonant acoustic field that causes them to levitate at mid-height and gather in two acoustic pressure nodes located on either side of the pre-focusing channel center, ***Fig. 1B***. The two bands of pre-focused cells enter the separation channel (30 mm × 380 μm × 150 μm) through the side branches of a trifurcation inlet where the cells are laminated along the channel side walls by a cell-free medium that enters through the central branch. The cells are here exposed to a ∼2 MHz half-wavelength acoustic standing wave field, which focuses the cells toward the center of the channel during their passage through the separation channel. Size is the predominant feature for how cells move in the sound field and larger cells (CTCs, yellow) migrate faster toward the channel center than smaller cells (WBCs, white), ***Fig. 1C***. The fraction of cells that exit through the central outlet can be tuned by adjusting the amplitude across the piezo electric actuator. For further details of acoustic cell manipulation ***see supplementary note and supplemental Figure 1***.

**Figure 1.**
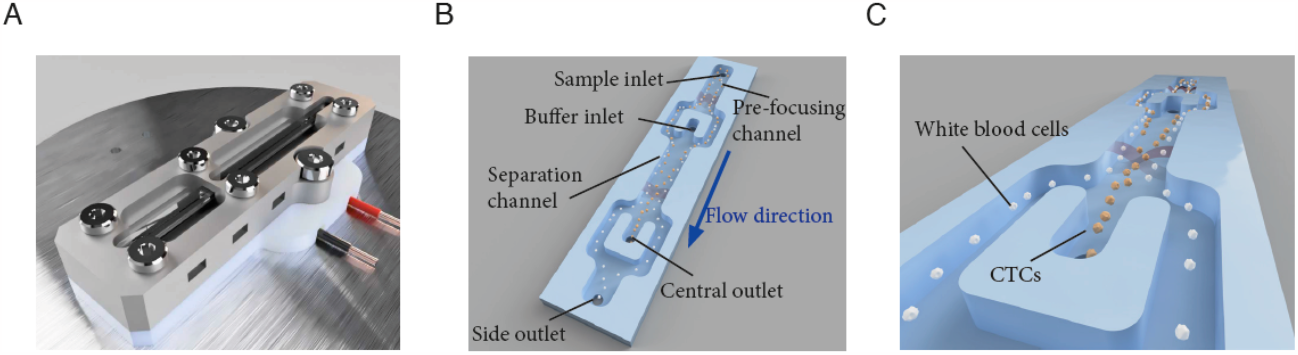
Illustration of the acoustofluidic microchip and the cell separation principle. A) The microfluidic chip. B) Key features of the microchip, C) Illustration of the acoustic focusing of cancer cells (yellow) towards the center of the separation channel and their following exit through the central outlet while WBCs (white) exit through the side-branches of the trifurcation outlet.

### Cell Culture

Human PCa cell lines DU145, PC3, LNCaP and breast cancer cell line MCF7 were acquired from American Type Culture Collection (ATCC). The cells were cultured according to recommendations at 37°C under a 5% CO2 atmosphere and harvested with trypsin-EDTA at approximately 80% confluency.

### Separation performance vs storage time

Cell lines, DU145, PC3, LNCaP and MCF7, were used for acoustophoretic cell separation from WBCs at 0, 1, 2 and 3 days post PFA fixation. The blood was subjected to RBC lysis using 1x BD FACS lysing solution according to manufacturer’s recommendations. All cells were fixed with a 4% PFA solution for 25 min in room temperature and stored at 4°C until the time of acoustic separation. Prior to separation, the cells were labeled with anti-EpCAM-PE and anti-CD45-APC for flow cytometry identification. Each sample was prepared with 0.05 mL RBC lysed blood diluted to a total sample volume of 0.2 mL and spiked with approximately 10,000 cancer cells. Three healthy blood donors were used for each cell line experiment, with six technical repeats for each time point. To account for biological differences of the cells on different days, and system variability, the acoustic energy was adjusted with the aim of retrieving above 90% of the cancer cells while keeping the WBC contamination well below 0.5%.

### Antibody panel, Immunostaining, Flow Cytometry enumeration, and IFC analysis

Cells were analyzed with a BD FACS CantoII or imaged by the image flow cytometer Amnis ImageStream Mk II (Millipore, Burlington, MA) and analyzed by the Ideas software. For CTC identification and WBC exclusion, acoustophoresis isolated cells were stained with a fluorescent antibody cocktail; anti-EpCAM-PE (BD biosciences) and anti-panCytokeratin (CK)-Alexa Fluor 488 (Thermo Fisher, Gothenburg Sweden), identifying CTCs, and anti-CD45-APC (BD biosciences) and anti-CD66b-Alexa Fluor 647 (BD biosciences) to distinguish WBCs. DAPI (Sigma-Aldrich) identified intact cells.

### Cell Search analysis

Isolation and enumeration of CTCs by CellSearch was performed at the Life Science Center, University of Düsseldorf, (Düsseldorf, Germany) following the manufacturer’s protocol. The CTC enumeration from acoustophoresis, using 6 mL of blood, was normalized to the blood volume, 7.5 mL, analyzed by the CellSearch assay when comparing the two technologies. Samples 11 and 12 are missing CellSearch data due to inability to analyze sample within 96 hrs. from blood draw.

### Data analysis

Cells classified as epithelial cells from the healthy control group was used to introduce a cut-off level of healthy men and men without metastases versus men with mPCa emitting CTCs to the circulation. The cutoff was chosen as the upper limit of the 99% confidence interval (i.e., mean + 2.6SD). To evaluate the chip’s ability to discriminate between cancer cell line cells and WBCs for increasing storage time, ***see supplemental note***.

## RESULTS AND DISCUSSION

### Separation outcome vs storage times

To establish a workflow from sample collection to cell characterization, we needed to measure the acoustophoresis separation efficiency over time. This followed the procedures reported in (26, 29). Mock samples with four cancer cell lines spiked in RBC-lysed blood were analyzed, but now extending the storage period over three days, and enumerating the proportion of cancer cells versus WBCs collected in the central outlet at different time points, ***Fig. 2A and Supplemental Fig. 2***. To determine whether the cell separation efficiency was significantly impacted by time elapsed between collection and acoustophoresis, we determined the deviation in the central fraction, fc, for all samples, ***Fig. 2B-C***. A linear fit shows that on average there is small, significant deviation in fc for WBCs, with a slope of 2.2·10^−4^ standard deviation of day 0 (SD_0_) as reference, over three days, the shift of the mean was 0.08%, corresponding to 1.2 SD_0_, and the SD at day 3 increased to 2.3 SD_0_. An unpaired Student’s t-test shows that the only significant change in the mean is between day 0 and either day 1, 2 or 3 (p1 = 3.4·10^−5^, p2 = 1.6·10^−3^, p3 = 8.3·10^−5^) but not from day 1 and forward. The trend for the cell lines is significantly declining with a slope of -4.4·10^−3^ per day (CI95, -6.8·10^−3^ to -1.9·10^−3^). Over three days, the shift of the mean was -1.3% which corresponds to 0.97 SD_0_, and the SD at day per day (CI95, 9.7·10^−5^ to 3.5·10^4^). Using the 3 increased to 2.3 SD_0_. Again, the t-test indicates a significant change from day 0 to day 1, 2 or 3 (p1 = 9.9·10^−4^, p2 = 2.5·10^−5^, p3 = 1.2·10^−3^). The fact that the slopes for the two categories of cells have opposite trends and increasing dispersion indicates that the ability to discriminate cancer cells from WBCs declines with time regardless of how the system is tuned, and cell separation performance will always be optimal on the day of blood draw.

**Figure 2.**
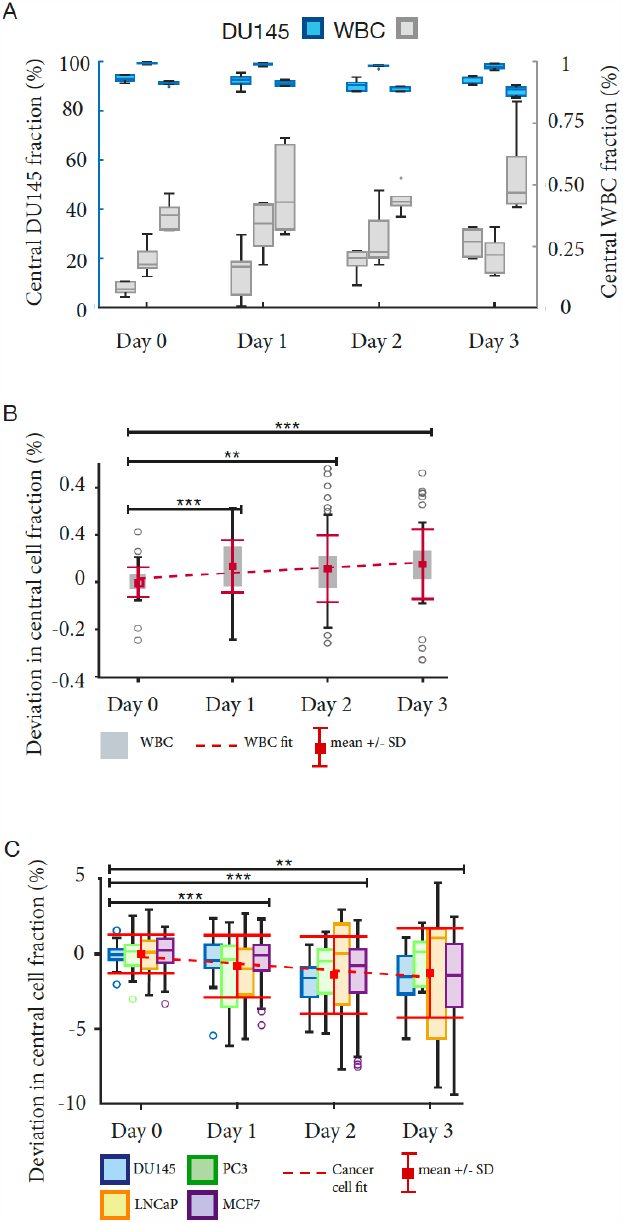
Separation performance for cancer cell line cells mixed with WBCs from healthy donors over time. (A) The central outlet fraction, fc, for DU145 and WBCs form healthy donors, (n=6). (B-C) Trend analysis of pooled data from (A) and Supplementary figure 1(A-C). Stars (*) indicate significance level relative to data from day 0.

### Antibody panel evaluation

The PCa cell line DU145 and RBC lysed blood from healthy donors were used as a model system to establish an antibody panel, (***Supplemental Fig. 3A-C)***. The autofluorescence of eosinophils in the green and yellow detection channels, [mainly due to flavin adenine dinucleotides (FAD)] (32), combined with low to undetectable expression of CD45 made it difficult to distinguish the eosinophils from cancer cells. The antibody anti-CD66b-AF647 (a granulocyte marker), was therefore added to the antibody panel, generating a strong signal in the red channel for all WBCs, identifying them as non-epithelial and thus excluding them from further analysis.

### Acoustophoretic CTC enrichment

The phenotypic definition of a CTC (EpCAM^+^, CK^+^, DAPI^+^ and CD45^-^), (***Fig. 3A panel I***), has been challenged. There is evidence that many CTCs have low or absent expression of epithelial markers after a phenotypic transformation during EMT (33). Label-free cell-separation using acoustophoresis manifest properties that could enable enrichment of CTCs with various lineage specificities. Therefore, we also used alternative CTC classifications to enumerate cells with little if any EpCAM expression (***Fig. 3A panel II***) or CK (***Fig. 3A panel III***), which cannot be enriched and detected using assay techniques employing EpCAM-antibody based isolation. However, further molecular characterization is needed to elucidate the origin and clinical value of these interesting cells. CTCs can occur in the blood both as single cells and cell clusters. The strong size dependency in acoustophoresis makes it particularly suitable for isolation of cell clusters, ***see supplemental note***. The interest in clusters has increased with reports on increasing levels of CTC-clusters with disease progression (34). We detected CTC-clusters (***Fig. 3B***), consisting of two or more cancer cells, as well as clusters of CTCs combined with various WBCs in most (9/12) of the analyzed mPCa cases. We enumerated CTCs (both single- and cluster-CTCs) from 6 mL of whole blood from 12 mPCa-cases. To maximize the recovery of small CTCs in the patient material, a background level of WBCs corresponding to 1-3% of the initial concentration in the sample was tolerated. This is higher compared to optimal settings for cell line separation. The contamination levels of WBC varied between the different patients due to variations in acoustic output level between experiments, and variations in WBC population in different patients. The major contaminants in the acoustic separation are eosinophils and other granulocytes (29), and samples with a high proportion of these cells will generate higher contamination levels.

**Figure 3.**
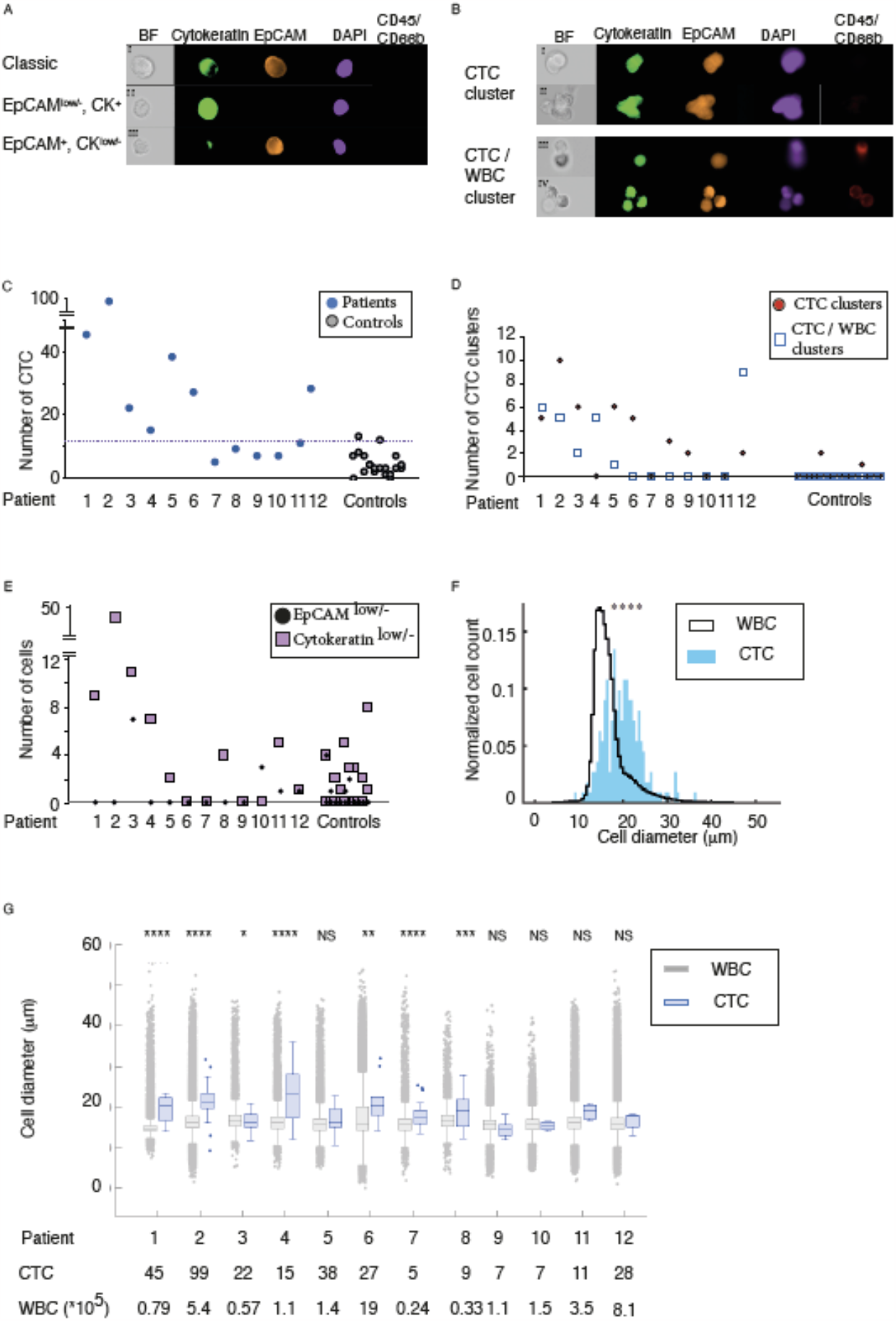
Circulating tumor cell (CTC) evaluation after acoustophoresis of blood from 12 metastatic prostate patients (A) CTCs and potential subtypes. (B) CTC Clusters (C) Enumeration of CTCs with EpCAM^+^,CK^+^, CD45^-^ and DAPI^+^ staining. (D) Number of detected cell clusters. (E) CTCs with low expression of EpCAM or Cytokeratin (F) Overall size and (G) Size distribution of CTCs and WBCs collected from the central outlet after acoustophoresis.

The acoustophoresis enriched CTCs were identified by epithelial phenotypic expression pattern (EpCAM^+^, CK^+^, CD45^-^, CD66b^-^, DAPI^+^) and morphological characteristics, ***Fig. 3C***. We calculated a threshold to discriminate enumeration of cells with epithelial phenotypic expression pattern in mPCa-cases compared to cells with epithelial expression found in healthy volunteers. Resulting in a cut-off of 13.8 cells (mean + 2.6SD) with epithelial phenotypic expression pattern in 6 mL of blood. When the threshold was rounded to nearest integer number of cells, there were 7 out of 12 mPCa cases displaying CTC numbers above this threshold. Using acoustophoresis, we detected a total of 67 CTC containing cell-clusters distributed among 9 out of 12 PCa-cases, ***Fig. 3D***. The clusters were divided into CTC-clusters consisting only of cancer cells (total 39 clusters) distributed among eight of the patients and CTC/WBC-clusters consisting of cancer cells aggregated to WBCs (total 28 clusters), distributed among 6 of the patients, ***Table 1***.. CTCs are known to associate with different cell types, such as neutrophils, fibroblasts, and platelets (14, 35-37) to form heterogenic clusters. The attachment of neutrophils has been hypothesized to support the metastatic potential of CTCs by amplifying their proliferative ability (14), while association with fibroblasts may enhance their metastatic capacity (35). Most detected cell clusters consisted of less than 4 associated cells, and two associated cells were the most common. Three cell clusters were also found among the 20 healthy control samples. These clusters consisted of two associated cells with epithelial expression pattern.

**Table 1.** Circulating tumor cell cluster distribution in metastatic prostate cancer patients.

| Patient # | CTC-clusters |  | CTC/WBC clusters |  |
| --- | --- | --- | --- | --- |
| | Number of cluster: with 2 CTCs | Cancer cell clusters with $\geq 3$ CTCs | Number of clusters: with 1 CTC | Number of clusters: with $\geq 2$ CTCs |
| 1 | 2 | 3 | 3 | 3 |
| 2 | 9 | 1 | 5 | 0 |
| 3 | 5 | 1 | 2 | 0 |
| 4 | 0 | 0 | 1 | 0 |
| 5 | 5 | 1 | 1 | 0 |
| 6 | 5 | 0 | 0 | 0 |
| 7 | 0 | 0 | 0 | 0 |
| 8 | 3 | 0 | 0 | 0 |
| 9 | 2 | 0 | 0 | 0 |
| 10 | 0 | 0 | 0 | 0 |
| 11 | 0 | 0 | 0 | 0 |
| 12 | 2 | 0 | 9 | 0 |

Our results agree well with previous reports indicating that CTC-clusters vary greatly in size, although smaller clusters between two and six cells are most common (34, 38). CellSearch did not detect any CTC-clusters in these patient samples. The analyzing program automatically discard pictures with CD45 positive cells, preclude detection of any heterogenous clusters containing both cancer cells and WBCs. The multitude of cell clusters found by acoustophoresis, may in addition to the acoustic-method’s sensitivity to size differences, be attributed to the relative gentleness of acoustic cell sorting (27). Moreover, putative CTCs with low EpCAM expression were found in four patients (No. 3, 10, 11 and 12). However equivalent cells could also be found in 20% of the control samples. In this study, patient number 3 was the only patient to exhibit CTCs with down-regulated EpCAM at a level above baseline, whereas three patients (No. 1-3) had elevated levels of cytokeratin low cells compared to healthy controls, ***Fig. 3E***. The current mode of acoustophoresis operation restricts isolation of smallest CTCs as their acoustophoretic mobility overlaps primarily with the granulocyte fraction, especially eosinophils. Hence, smaller CTCs may be lost in the WBC side fraction and due to the overwhelming WBC count, the fraction of missed CTCs have not been possible to establish. As acoustic separation is highly dependent on cell size, it carries an inherent risk of not detecting small CTCs, not diverted to the central outlet. The CTCs diverted to the central outlet by acoustophoresis were generally larger than contaminating WBCs and varied in size between 7 and 28 μm in diameter, (median size: 17.0 μm). There was no apparent correlation between cell size and detected number of CTCs in blood from mPCa-cases, ***Fig. 3F-G***. On-going developments are aiming at reducing the acoustophoretic processing time by increasing the acoustic energy density in the pre-focusing and the separation channel by optimizing the acoustic resonance conditions of the chip design. An optional method for improvement was introduced by Augustsson et al. (39), demonstrating an acoustophoresis modality where the use of a buffer density gradient makes the acoustophoretic separation independent of cell size, though currently at a modest throughput. An analogous acoustophoresis approach that more easily could enable high-throughput processing would be to use buffer density step gradients defined to discriminate specific cell populations (40). Possible further improvements encompass optimization of side and center input and output split flow ratio, matched to flow rate, and input acoustic power.

### Enumeration of CTCs after acoustophoresis compared to CellSearch

We evaluated the performance of acoustophoresis with 6 mL EDTA-anti-coagulated blood from ten mPCa-cases who also provided 7.5 mL of CellSave blood at the same venipuncture for CellSearch analysis. Overall, the number of CTCs detected after acoustophoresis were higher compared to CellSearch in all mPCa-cases ***Fig. 4***. However, the detection of an average of six clusters in 6 mL blood from 8/10 mPCa-cases was unique to acoustophoresis as no CTC-clusters were detected in the corresponding blood sample analyzed by CellSearch. Interestingly, one patient with mPCa had high number of CTCs detected after acoustophoresis and only one CTC detected by CellSearch and prostate specific antigen (PSA) level < 1 ng/mL in serum. The CTCs detected in this patient had the largest measured mean size, with substantive variation in size distribution among CTCs detected.

**Figure 4.**
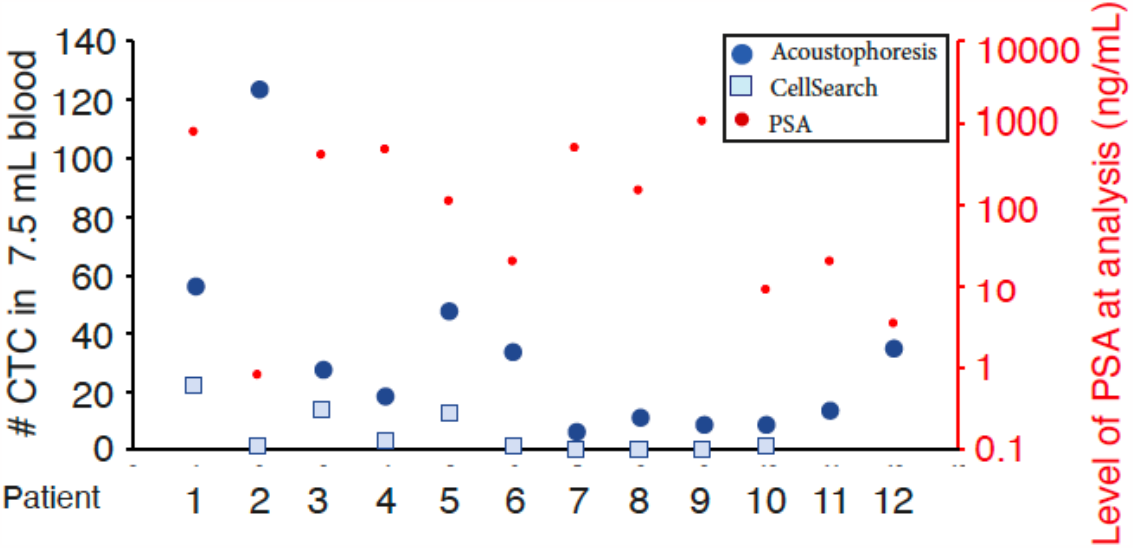
Comparison of Circulating tumor cell enumeration for acoustophoresis and CellSearch. CTCs with EpCAM^+^,CK^+^, CD45^-^ and DAPI^+^ staining were enumerated for 7.5 mL blood from 12 patients with metastatic prostate cancer with acoustophoresis (blue circles) and patients 1-10 with CellSearch (squares), correlated to PSA value at time of sample analysis (red circles).

## Conclusion

Acoustophoresis holds promise as an inclusive CTC technology due to its label-free separation approach which allows for the isolation of various CTC subtypes, including cells undergoing EMT. Due to the gentle separation approach, it is also effective in isolating CTC-clusters which is suggested as a marker for aggressive disease. To confirm the value of acoustophoresis we performed a comparison versus the CellSearch system. To create a more comprehensive picture of the CTC population in a patient, it is vital to include the wider spectrum of inclusion criteria offered by acoustophoresis, including CTC-clusters, and different subclasses of CTCs with altered expression patterns compared to the classic CTC profile. The increased levels of detected CTCs with acoustophoresis offer promise for future detection and characterization of CTCs earlier in the disease progression. Even more promising is the detection of several CTC-clusters in the patients with metastatic disease, which was not detected in healthy controls. However, for a more conclusive evaluation of the developed methodology a future validation study including a larger cohort of patient samples and healthy controls is required.

## Data Availability

All data produced in the present study are available upon reasonable request to the authors.

## ASSOCIATED CONTENT

### Supporting Information

Supplemental note describing Acoustic cell separation theory and cell movement as well as data analysis (pdf).

Supplemental Table 1, with patient information (pdf)

Supplemental figures 1-3 describing the acoustophoretic principle, additional separation performances and antibody control staining (pdf)

## AUTHOR INFORMATION

### Author Contributions

The manuscript was written through contributions of all authors. All authors have given approval to the final version of the manuscript.

## ACKNOWLEDGMENT

The CellSearch CTC isolation and evaluations were performed at the Life Science Center at the University of Düsseldorf, (Düsseldorf, Germany).

