## Supplementary material for "Acoustic enrichment of heterogenous circulating tumor cells and clusters from patients with metastatic prostate cancer"

**Supplementary Table 1.** Prostate cancer patient information overview

| Patient # | 1 | 2 | 3 | 4 | 5 | 6 | 7 | 8 | 9 | 10 | 11 |
|---|---|---|---|---|---|---|---|---|---|---|---|
| Hospital site | Gothenburg | Malmö | Malmö | Malmö | Malmö | Malmö | Gothenburg | Gothenburg | Malmö | Malmö | Malmö |
| Date of diagnosis | Oct-19 | Aug-21 | Jan-22 | Apr-22 | Jun-22 | Sep-22 | Mar-18 | Jan-20 | Oct-22 | Oct-22 | Aug-21 |
| Date of sample analysis | Sep-20 | Nov-21 | Mar-22 | May-22 | Sep-22 | Nov-22 | Nov-22 | Nov-22 | Nov-22 | Nov-22 | Nov-21 |
| Age range at analysis (Years) | 90-94 | 75-79 | 70-74 | 75-80 | 65-69 | 55-59 | 75-79 | 75-79 | 90-94 | 80-84 | 80-84 |
| TNM | T4NxM1 | T3T4N1M1 | T4N1M1 | T2N1M1 | T4N1M1 | T3N1M1 | TxNxM1 | T3NxM1 | TXN1M1 | T3N1M1 | T3N1M1 |
| Gleason score | 4+5 | 4+5 | 4+5 | N.A. | 4+5 | 4+5 | 4+4 | N.A. | N.A. | 4+5 | 4+5 |
| Location of metastasis | bone | LN, bone | LN, bone | LN, bone | LN, bone | LN, bone, lung | bone | bone | LN, bone | LN, bone | LN, bone |
| PSA level sample analysis (ng/mL) | 770 | 0.81 | 397 | 476 | 112 | 20 | 490 | 150 | 1015 | 9.0 | 20 |
| PSA level diagnosis (ng/mL) | 1100 | 60 | 3518 | 6647 | 118 | 312 | 170 | 1700 | 1015 | 989 | 26* |
| Alkaline phosphatase level | 19.3 | 2.7 | 38 | 8.7 | 1.5 | 2.0 | 1.6 | 11 | 3.4 | 12 | 1.9 |
| Treatment | GnRH** | GnRH** | GnRH** | GnRH** | GnRH** Bikalutamid | GnRH** | GnRH** *** | GnRH** **** | GnRH** | GnRH** Bikalutamid Abiraterone | GnRH** Bikalutamid |
| CellSearch | Y | Y | Y | Y | Y | Y | Y | Y | Y | Y | N |

* PSA level 2 months prior to diagnosis
** Any GnRH-analogue, in som patients preceeded by one or two months GnRH-antagonist
*** Patient 7 recieved also other treatments, but not at the time of sample analysis (Docetaxel, Enzalutamide, Radium).
**** Patient 8 recieved also other treatments, but not at the time of sample analysis (Abiraterone).
LN = lymphnode

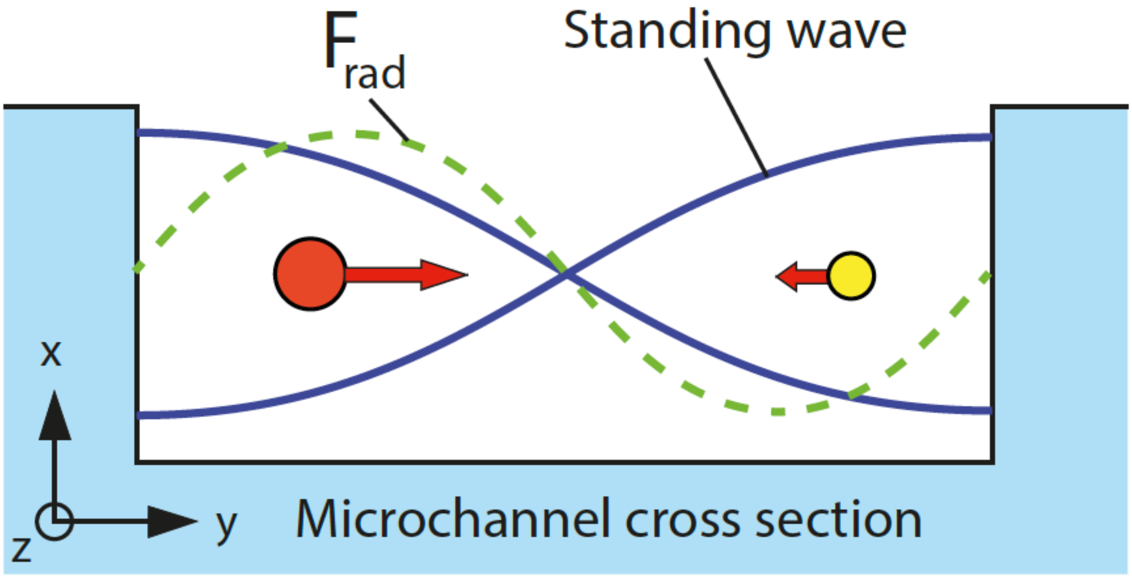


***Supplemental Figure 1. Schematic of the acoustophoresis principle,*** where cells with a higher acoustophoretic mobility migrates faster towards the standing wave pressure node in the center of the channel while following the flow in the z-direction to the outlets.

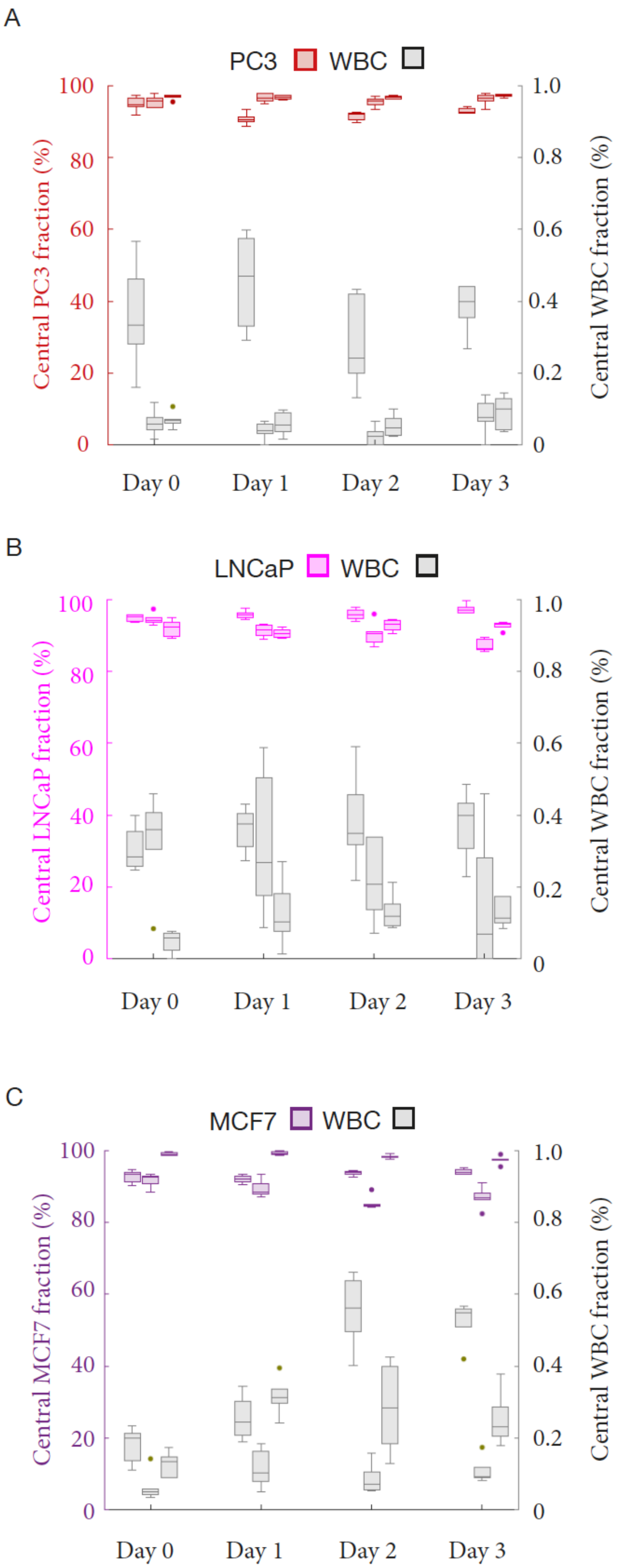

A
PC3
WBC
Central PC3 fraction (%)
Central WBC fraction (%)
100
80
60
40
20
0
1.0
0.8
0.6
0.4
0.2
0
Day 0
Day 1
Day 2
Day 3
B
LNCaP
WBC
Central LNCaP fraction (%)
Central WBC fraction (%)
100
80
60
40
20
0
1.0
0.8
0.6
0.4
0.2
0
Day 0
Day 1
Day 2
Day 3
C
MCF7
WBC
Central MCF7 fraction (%)
Central WBC fraction (%)
100
80
60
40
20
0
1.0
0.8
0.6
0.4
0.2
0
Day 0
Day 1
Day 2
Day 3

***Supplemental Figure 2. Acoustic separation performance for cancer cell line cells mixed with WBCs from healthy donors over time.*** (A)PC3, (B) LNCAP, and (C) MCF7. The horizontal line inside each box represents the median, top and bottom edges represent the upper and lower quartiles, whiskers indicate non-outlier minimum and maximum, and rings indicates values outside 1.5 times the interquartile range, (n=6).

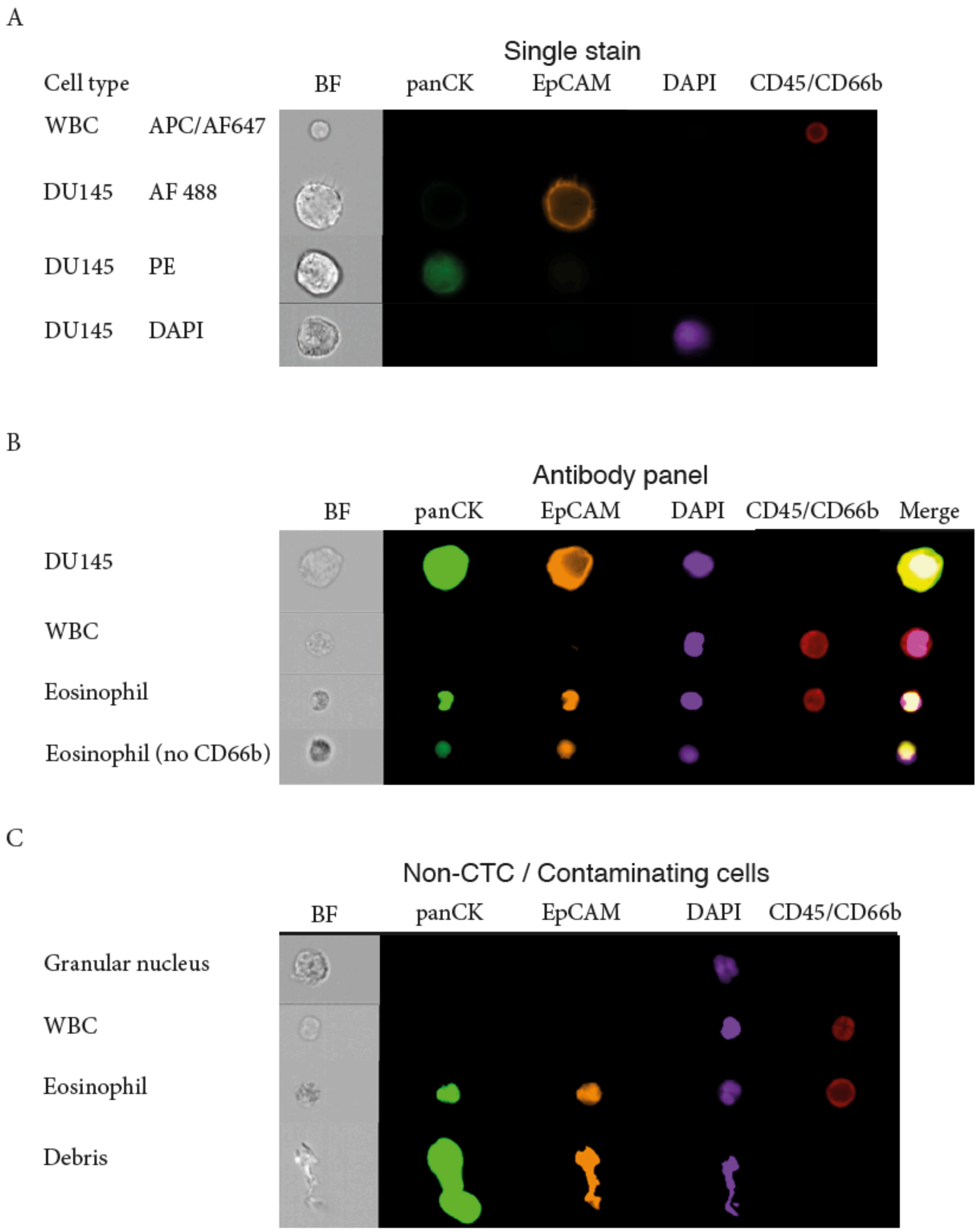


***Supplemental Figure 3. Control staining of DU145 cell line and WBCs from a healthy blood donor and contaminating non-CTC cells from mPCa patient samples.*** (A) Single antibody staining of control cells (B) Representative images using the full antibody panel. (C) Examples of representative contaminating cells found in the central fraction from patient #1, which were discarded as non-CTCs.

***Supplementary note 1:***

***Theory behind acoustic cell separation:***

Acoustophoresis utilizes the difference in acoustophoretic mobility between cell species, i.e., the acoustic radiation force, $F_{rad}$ (eq. 1), acting on cells depending on the cells specific acoustophysical properties, i.e., size, density, and compressibility. Cells exposed to an acoustic standing wave, as illustrated in Fig 1B and C, will migrate to the center of the microchannel at a velocity, $u_y$ (eq. 2), determined by its physical properties while following the flow along the channel. Cells that have a higher acoustophoretic mobility will migrate faster to the channel center and are collected at the central outlet, which is the case for the CTCs in this study. Slower migrating cells, predominantly WBCs will be discarded via the side outlet.

Equation (1) shows the position ($y_c$) dependent force ($F_{\text{rad}}$) acting on a suspended cell of radius ($a$), density $\rho_c$, and compressibility $\kappa_c$ in a plane standing wave of acoustic energy density $E_{\text{ac}}$ with a pressure node located at $y = 0$ [1].

$$F_{\text{rad}} = -4E_{\text{ac}}\, k_y\, \pi\, a^3 \Phi \sin(2k_y\, y_p) \tag{1a}$$

$$\Phi = \frac{1-\tilde{\kappa}}{3} + \frac{\tilde{\rho}-1}{2\tilde{\rho}+1} \tag{1b}$$

Here $k_y = 2\pi/\lambda$ is the wave vector and $\tilde{\kappa} = \kappa_c/\kappa_m$ and $\tilde{\rho} = \rho_c/\rho_m$ is the relative compressibility and density of the cell with respect to the suspending medium (indicated with superscript *m*).

The resulting velocity of the cell due to drag from the fluid is expressed in equation (2).

$$u_y = F_{\text{rad}} \,/\, 6\pi\eta a \propto \Phi a^2 \tag{2}$$

The velocity of a cell is thus proportional to the square of the cell radius, which explains the strong dependency of the cell size, while differences in density and compressibility lead to a linear change.

Supplemental figure 1 schematically shows the cross section of a microchannel where a CTC and a WBC are exposed to the acoustic radiation force, resulting in a higher migration velocity (red arrow) to the channel center as compared to WBCs. Direction of buffer flow is in the z-direction.

***Data analysis of discrimination between cancer cell line cells and white blood cells for increasing storage time in the acoustophoretic CTC chip:***

To discriminate between cancer cell line cells and WBCs for increasing storage time, the number of cells of each type in the central ($N_c$) and side ($N_s$) outlet streams were counted after each run. The central fraction of a cell type ($i$) was calculated as the relative fraction ($f_{c,i}$) of that cell in the center *vs* all retrieved cells, *i.e.*, $f_{c,i} = N_{c,i} / ( N_{c,i} + N_{s,i})$. The ideal separation outcome is thus $f_c$ = 100% for cancer cells and $f_c$ = 0% for WBCs.

To measure the deviation of $f_c$ *vs* storage time in a donor specific manner, we computed the mean for the six technical repeats on day 0, $\langle f_c(0) \rangle$, for each of the three donors, and subtracted that from $f_c(t)$ for all the different storage times. The drift vs storage time was then estimated as the slope, with 95% confidence interval (CI95), of $f_c(t) - \langle f_c(0) \rangle$ after linear fitting.

### *Cell cluster movement during acoustophoresis:*

The strong size dependency for cell movement in acoustophoresis makes this method particularly suitable for isolation of cell clusters. For a cluster of $N$ cells, the acoustic migration velocity $u_{\text{ac}}$ is expected to scale with $N^{2/3}$ such that even a cluster of just two cells will move towards the center at ~1.6 times the velocity of a single cell. Here we have assumed that (*i*) the force on the cluster increases with the total volume $V_{\text{cluster}} = N \cdot V_{\text{cell}}$, (*ii*) the shape of the cluster is approximated as a sphere of volume $V_{\text{cluster}}$, and (iii) the hydrodynamic drag force scales with the radius of a sphere.
